# Expansion of Cytotoxic CD4+ T cells in the lungs in severe COVID-19

**DOI:** 10.1101/2021.03.23.21253885

**Authors:** Naoki Kaneko, Julie Boucau, Hsiao-Hsuan Kuo, Cory Perugino, Vinay S. Mahajan, Jocelyn R. Farmer, Hang Liu, Thomas J. Diefenbach, Alicja Piechocka-Trocha, Kristina Lefteri, Michael T. Waring, Katherine R. Premo, Bruce D. Walker, Jonathan Z. Li, Gaurav Gaiha, Xu G. Yu, Mathias Lichterfeld, Robert F. Padera, Shiv Pillai

**Author notes:** Correspondence to (SP) and (RFP).

## Abstract

The contributions of T cells infiltrating the lungs to SARS-CoV-2 clearance and disease progression are poorly understood. Although studies of CD8+ T cells in bronchoalveolar lavage and blood have suggested that these cells are exhausted in severe COVID-19, CD4+ T cells have not been systematically interrogated within the lung parenchyma. We establish here that cytotoxic CD4+ T cells (CD4+CTLs) are prominently expanded in the COVID-19 lung infiltrate. CD4+CTL numbers in the lung increase with disease severity and progression is accompanied by widespread HLA-DR expression on lung epithelial and endothelial cells, increased apoptosis of epithelial cells and tissue remodeling. Based on quantitative evidence for re-activation in the lung milieu, CD4+ CTLs are as likely to drive viral clearance as CD8+ T cells and may also be contributors to lung inflammation and eventually to fibrosis in severe COVID-19.

**Graphical Abstract:** 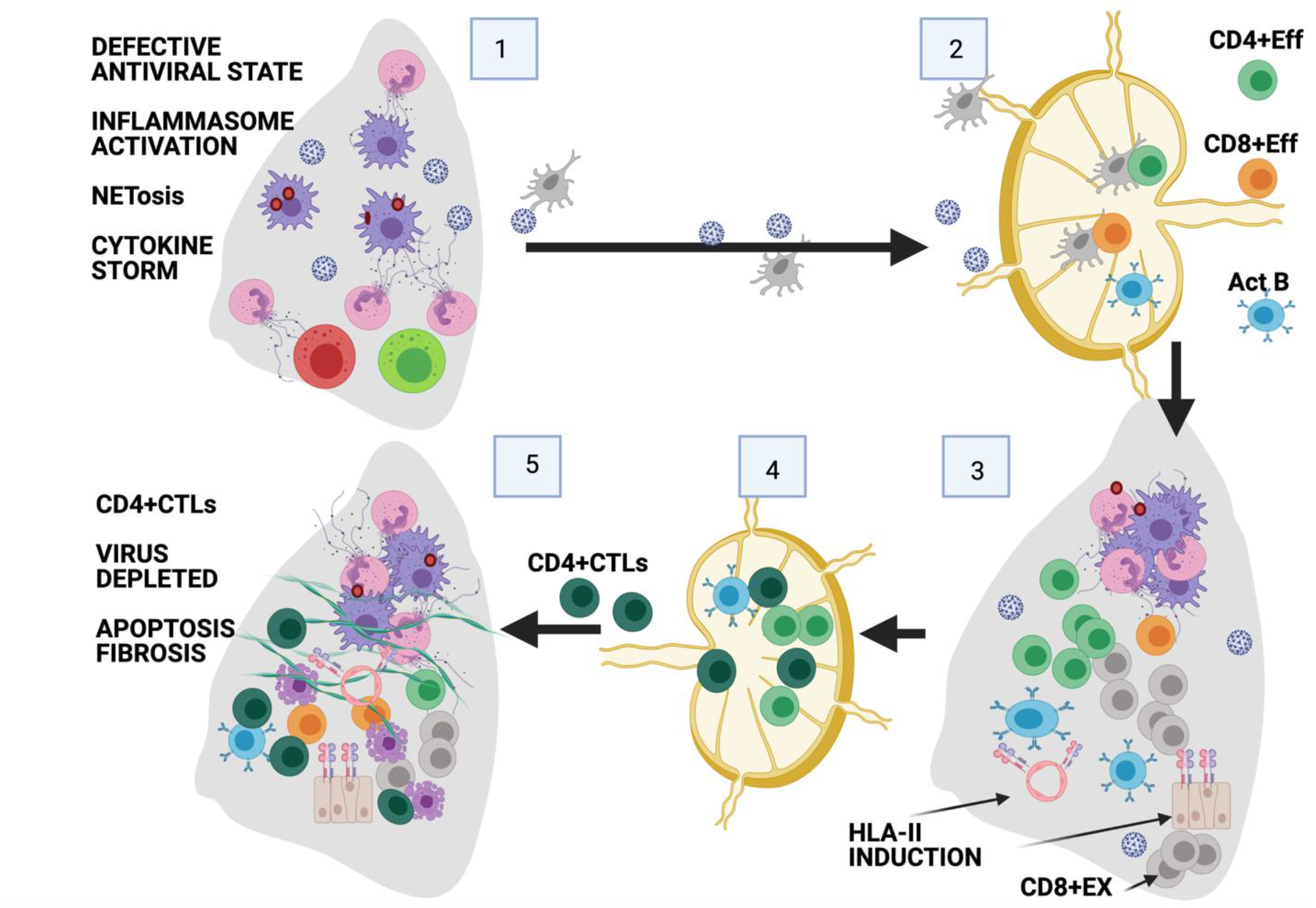

**In Brief:** In severe COVID-19 cytotoxic CD4+ T cells accumulate in draining lymph nodes and in the lungs during the resolving phase of the disease. Re-activated cytotoxic CD4+ T cells and cytotoxic CD8+ T cells are present in roughly equivalent numbers in the lungs at this stage and these cells likely collaborate to eliminate virally infected cells and potentially induce fibrosis. A large fraction of epithelial and endothelial cells in the lung express HLA class II in COVID-19 and there is temporal convergence between CD4+CTL accumulation and apoptosis in the lung.

**Highlights:**

- In severe COVID-19, activated CD4+ CTLs accumulate in the lungs late in disease
- These cells likely participate in SARS-CoV-2 clearance, collaborating with CD8+ T cells many of which exhibit an exhausted phenotype
- T cells likely contribute to the late exacerbation of inflammation
- CD4+CTLs have been linked to fibrosis in many disorders and could also be responsible for the eventual induction of fibrosis in a subset of COVID-19 patients

**Summary:** The contributions of T cells infiltrating the lungs to SARS-CoV-2 clearance and disease progression are poorly understood. Although studies of CD8+ T cells in bronchoalveolar lavage and blood have suggested that these cells are exhausted in severe COVID-19, CD4+ T cells have not been systematically interrogated within the lung parenchyma. We establish here that cytotoxic CD4+ T cells (CD4+CTLs) are prominently expanded in the COVID-19 lung infiltrate. CD4+CTL numbers in the lung increase with disease severity and progression is accompanied by widespread HLA-DR expression on lung epithelial and endothelial cells, increased apoptosis of epithelial cells and tissue remodeling. Based on quantitative evidence for re-activation in the lung milieu, CD4+ CTLs are as likely to drive viral clearance as CD8+ T cells and may also be contributors to lung inflammation and eventually to fibrosis in severe COVID-19.

## Introduction

Susceptibility to severe COVID-19 is best understood in the context of genetic defects in the Type I interferon pathway or the presence of autoantibodies to Type I interferons (Zhang et al., 2020a; Bastard et al., 2020). The attenuation of the production of, or responses to, Type I interferons likely allows SARS-CoV-2 to replicate more efficiently and establish itself in the lungs. Other factors such as male gender, obesity and advanced age also contribute but the mechanisms that underlie their contributions to susceptibility are less well understood. The accumulation of virus in the respiratory tract and the resultant cell damage and/or cell death results in the over-exuberant activation of neutrophils, monocytes and macrophages by PAMPs and DAMPs leading to the development of acute respiratory distress syndrome, cytokine storm and progressive lung damage.

Much less is understood regarding the pathogenesis of severe COVID-19 once the virus begins to replicate in the lungs. Our previous studies revealed that in acutely ill patients who succumbed in less than ten days after the onset of symptoms, SARS-CoV-2 was readily detectable in the lungs, but in patients with resolving disease, very little virus was detectable in the lungs (Schaefer et al., 2020). Although clearance of virus from the lungs typically occurs during the resolving phase of infection in severely ill patients, no specific T cell subset has been firmly implicated in this process, especially since there are varying reports, none directly examining the lung, regarding the potential loss of functionality of CD8+ T cells in severe disease. Very little is also known regarding the potential contribution of adaptive immune cells to progressive lung damage and the triggering of fibrosis in severe COVID-19.

Although numerous studies have been performed on circulating T cell subsets in severe COVID-19, there have been technical challenges to quantitating T cell subsets that infiltrate the diseased lung using flow cytometry, mass cytometry and single-cell transcriptomic approaches. As a result, T cell subsets at the major site of inflammation and tissue damage have so far not been systematically qualitatively or quantitatively categorized in this disease.

Here we describe the quantitative analysis of innate and adaptive immune cells in the lungs and draining thoracic lymph nodes in patients with acute and resolving severe COVID-19. Our studies show that inflammatory myeloid cells continue to infiltrate the lung and be activated as disease progresses. In contrast, innate lymphoid cells that are generally implicated in viral clearance, namely NK cells and ILC1 cells, were not prominent in the lungs during severe disease. Late in severely ill COVID-19 patients, some CD8+ T cells exhibited signs of recent activation, but most CD8+ T cells in the lung express markers of exhaustion.

Among CD4+ T cell subsets, cytotoxic CD4+ T cells or CD4+CTLs were prominent in the lung and increased in absolute numbers and frequency in the resolving phase of the disease. Many of these lung-infiltrating CD4+ T cells expressed Granzyme B and their presence in the lungs was accompanied by HLA-DR expression in a large proportion of epithelial and endothelial cells in these inflamed organs. A major increase in apoptosis of epithelial cells was observed late in the disease at a time when CD4+CTLs had expanded, (and SARS-CoV-2 had also been depleted). CD4+CTLs expand in draining lymph nodes, infiltrate the lungs and are likely to be as relevant to viral clearance as CD8+ cytotoxic cells in this disease. We show here that CD4+CTLs also expand in the blood as disease severity increases. CD4+CTLs are among the few antigen-specific CD4+ T cell subsets that have been reported to be expanded in the blood of severely ill COVID-19 patients (Meckiff et al. 2020) as determined by single cell RNA-sequencing, consistent with our observations at the primary site of severe disease.

CD8+ T cell exhaustion is thought to have evolved to limit organ damage in severe viral infections (Zajac et al., 1998; Cornberg at al., 2013; Wherry and Kurachi, 2015). CD4+CTLs may have in turn evolved to more selectively eliminate viruses in the context of inflammation and possibly of co-existing CD8+ T cell exhaustion.

Given the known links of CD4+CTLs to fibrosis in a number of diseases (Mattoo et al., 2016; Maehara et al., 2020; Perugino et al., 2021; Allard-Chamard et al., 2021; Wang et al., 2021), these T cells may be key contributors to the acceleration of apoptotic death, inflammation and possibly the eventual induction of lung fibrosis already documented in severe COVID-19.

## Results

### Inflammation increases late in severe COVID-19 even after viral depletion

We quantitatively analyzed lungs and thoracic lymph nodes from two groups of individuals who had succumbed to severe COVID-19. The first or “early” group had had disease for a shorter duration (under 10 days from the onset of symptoms) and represents a group in which SARS-CoV-2 is abundant in the lungs (Schaefer et al., 2020), whereas the second “late” or resolving group had had disease for 15-36 days, a time at which virus is more difficult to detect (Table S1). Staining of epithelial and endothelial cells confirmed that lung architecture is severely distorted in COVID-19 (Figure 1A) but infiltrating neutrophils (frequently NETosing, as revealed by citrullinated histone H3) and macrophages (with frequent inflammasome related ASC-1 specks) were even more abundant late in the disease, indicative of ongoing inflammation at a time when virus had largely been eliminated from the lung (Figures 1B to 1E). Although NK cells and ILC1s can theoretically participate in viral clearance they were not prominent in early or late patients and total ILCs in the lung were also largely unchanged (Figures 1F and G, Figure S1).

**Figure 1.**
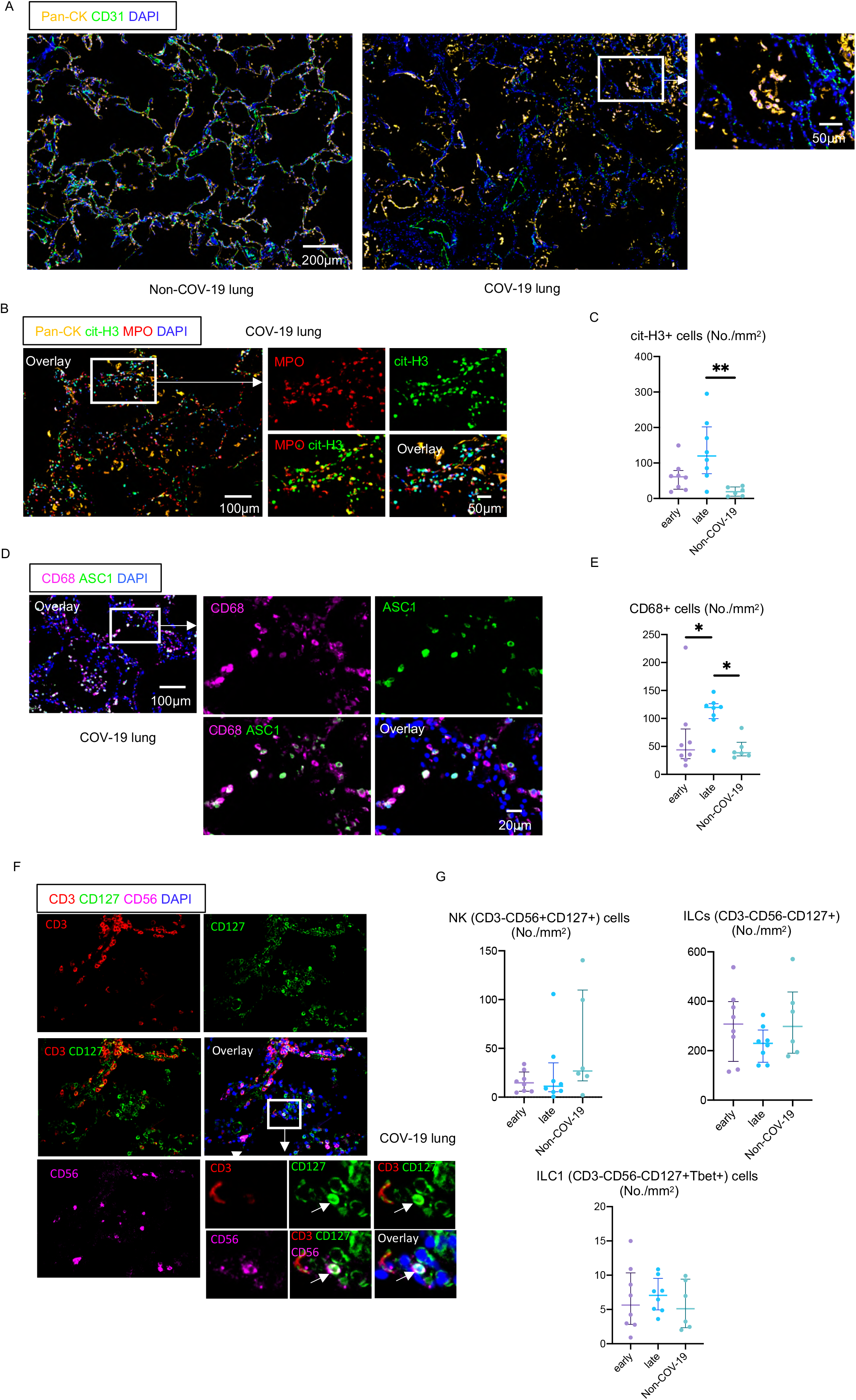
Macrophages and neutrophils dominate the lung inflammatory phenotype but NK cells and ILCs are not prominent in COVID-19 lungs. (A) Representative multi-color immunofluorescence images of Pan-Cytokeratin (Pan-CK; orange), CD31 (green) and DAPI (blue) staining of lung sections from non-COVID-19 (Non-COV-19) and COVID-19 (COV-19) patients (left and middle). Right panel showing desquamation of Pan-CK+ epithelial cells into the alveolar spaces. (B) Representative multi-color immunofluorescence images of Pan-CK (orange), citrullinated histone 3 (cit-H3) (green), myeloperoxidase (MPO) (red) and DAPI (blue) staining in a lung from a COVID-19 patient. (C) Absolute numbers of cit-H3+ cells in lungs from early (purple) (n = 8) and late (blue) (n = 8) COVID-19 patients and non-COVID-19 patients (green) (n = 6). (D) Representative multi-color immunofluorescence images of CD68 (purple), ASC1 (green) and DAPI (blue) staining in a lung from a COVID-19 patient. (E) Absolute numbers of CD68+ cells in lungs from early (purple) (n = 8) and late (blue) (n = 8) COVID-19 patients and non-COVID-19 patients (green) (n = 6). (F) Representative multi-color immunofluorescence images of CD3 (red), CD127 (green), CD56 (purple) and DAPI (blue) staining in a lung from a COVID-19 patient. Arrow indicates a CD3-CD127+ CD56+ NK cell. (G) Absolute numbers of NK cells (upper left), ILCs (upper right), ILC1s (lower middle) in lungs from early (purple) (n = 8) and late (blue) (n = 8) COVID-19 patients and non-COVID-19 patients (green) (n = 6). All p-values were calculated using the Kruskal-Wallis test to control for multiple comparisons. *p < 0.05; **p < 0.01.

### Increased infiltration of the lungs by CD4+CTLs is prominent as severe COVID-19 progresses

We systematically quantitated prominent CD4+ T cell subsets in patients with COVID-19 in the lung and in draining thoracic lymph nodes. In spite of documented CD4+ T cell lymphopenia in the lymph nodes in severe COVID-19, (Kaneko et al., 2020; see also Figure 3), the infiltration of the lungs by CD4+T cells was comparable in absolute numbers with that seen in lung inflammation caused by non-COVID-19 linked causes and trended towards an increase in patients who had had COVID-19 for a longer period of time. However, the absolute numbers and relative proportions of T-bet expressing T_H_1 cells, GATA-3 expressing T_H_2 cells, RORC expressing T_H_17 cells, CXCR5 expressing T_FH_ cells and FOXP3 expressing T_regs_ in the lungs in COVID-19 were comparable to the numbers seen in patients with non-COVID-19 linked lung disease (Figure 2 and Figures S2A and B)

**Figure 2.**
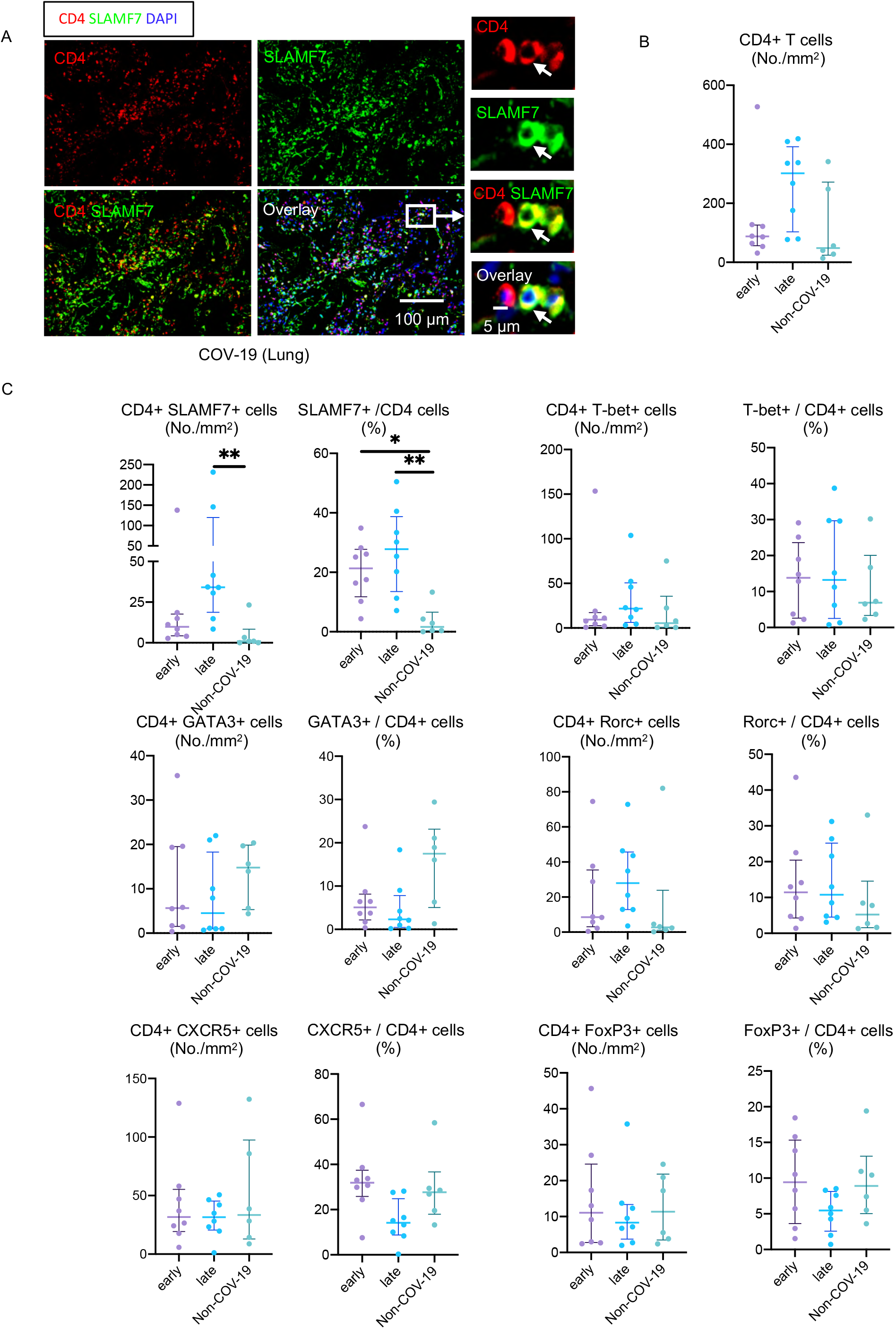
CD4+CTLs represent an expanded CD4+ T cell population in the lungs of COVID-19 patients. (A) Representative multi-color immunofluorescence images of CD4 (red), SLAMF7 (green) and DAPI (blue) staining in a lung from a COVID-19 patient. Arrows indicate a. CD4+ SLAMF7+ CD4+CTL. (B) Absolute numbers of total CD4+ T cells in lungs from early (purple) (n = 8) and late (blue) (n = 8) COVID-19 patients and non-COVID-19 patients (green) (n = 6). (C) Absolute numbers and relative proportions of CD4+CTLs (upper left), T_H_1 (upper right), T_H_2 (middle left), T_H_17 (middle right), T_FH_ (lower left) and T_reg_ (lower right) cells in lungs from early (purple) (n = 8) and late (blue) (n = 8) COVID-19 patients and non-COVID-19 patients (green) (n = 6). All p-values were calculated using the Kruskal-Wallis test to control for multiple comparisons. *p < 0.05; **p < 0.01.

**Figure 3.**
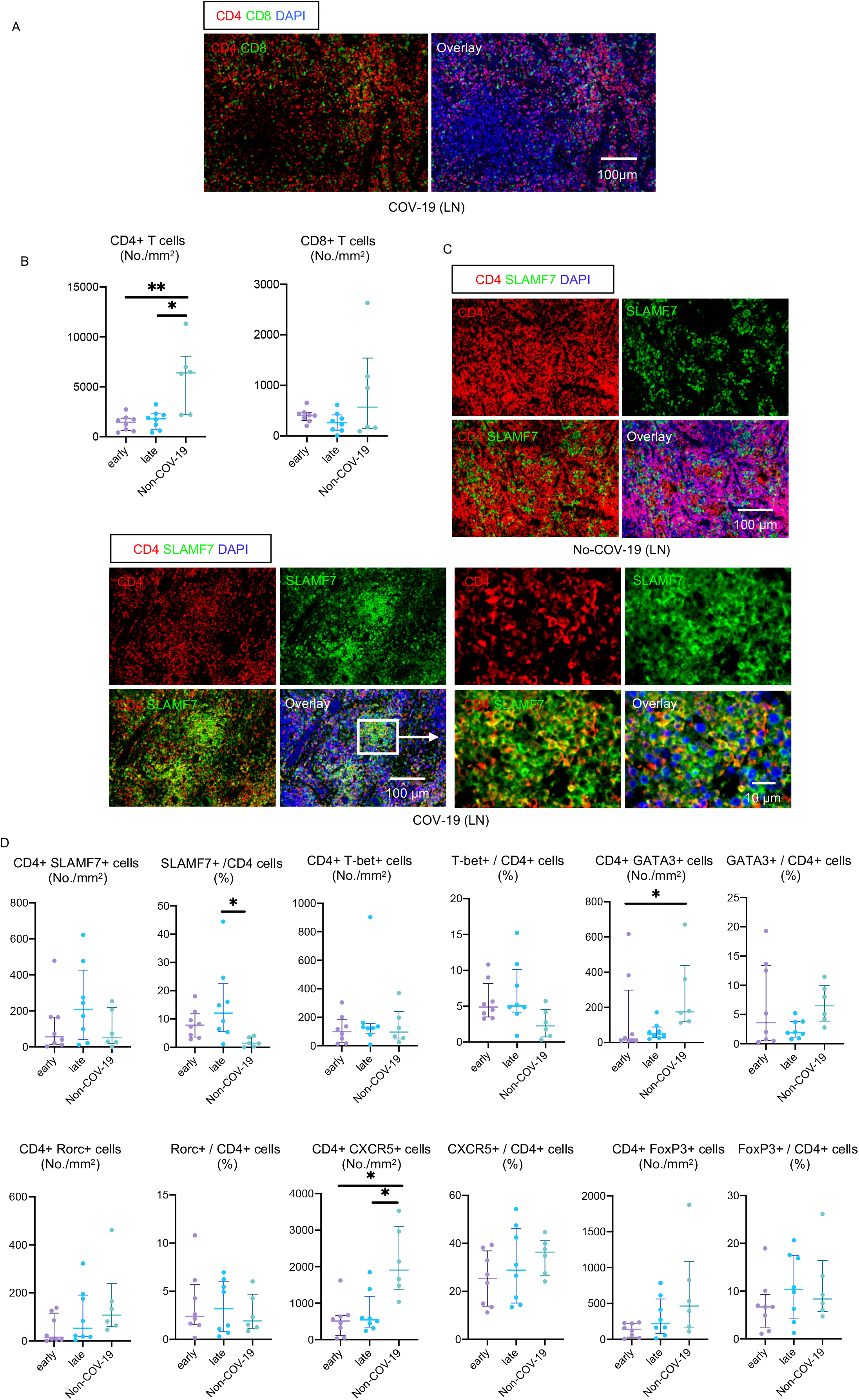
Relative increase in CD4+CTLs but no expansion of CD8+ T cells in COVID-19 lymph nodes. (A) Representative multi-color immunofluorescence images of CD4 (red), CD8 (green) and DAPI (blue) staining in a lymph node from COV-19 patient. (B) Absolute numbers of total CD4+ T cells (left) and CD8+ T cells (right) in lungs from early (purple) (n = 8) and late (blue) (n = 8) COVID-19 patients and non-COVID-19 patients (green) (n = 6). (C) Representative multi-color immunofluorescence images of CD4 (red), SLAMF7 (green) and DAPI (blue) staining in lungs from non-COVID-19 patient (upper) and a COVID-19 patient (lower). Yellow cells with red (CD4) and green (SLAMF7) merged are CD4+CTLs and are frequently seen in COVID-19 patients. In contrast, red and green can still be separately identified, and yellow cells are relatively sparse in non-COVID-19 patients. (D) Absolute numbers of total CD4+ T cells in lymph nodes from early (purple) (n = 8) and late (blue) (n = 8) COVID-19 patients and non-COVID-19 patients (green) (n = 6). (C) Absolute numbers and relative proportions of CD4+CTLs (upper left), T_H_1 (upper middle), T_H_2 (upper right), T_H_17 (lower left), T_FH_ (lower middle) and T _reg_ (lower right) cells in lungs from early (purple) (n = 8) and late (blue) (n = 8) COVID-19 patients and non-COVID-19 patients (green) (n = 6). All p-values were calculated using the Kruskal-Wallis test to control for multiple comparisons. *p < 0.05; **p < 0.01

The one CD4+ T cell subset that was clearly expanded in the lungs of COVID-19 patients was the CD4+SLAMF7+ CD4+CTL population (Figures 2A and B). A very similar increase was observed using CX3CR1 instead of SLAMF7 as a marker for CD4+CTLs (Figures S2C and D) (Weiskopf et al., 2015, Perugino et al., 2021). CD4+CTLs were increased both in terms of absolute numbers as well as proportions and this increase was most significant in late or resolving patients.

Examination of the draining lymph nodes revealed CD4+ T cell lymphopenia and a late increase in CD4+CTLs (Figure 3 and Figure S2E). The increase in CD4+CTLs in draining lymph nodes was also validated by CX3CR1 staining (Figure S2F and G). Relative attenuation was seen of T_H_2 cells and CXCR5+CD4+ T_FH_ cells in the lymph nodes. Overall, these data indicated that CD4+CTLs represent a prominent CD4+ T cell subset that infiltrates and/or expands in the major affected organ in COVID-19, and this infiltration/expansion is most prominent late in the disease (in spite of CD4+ T cell lymphopenia in the draining lymph nodes). No expansion of CD8+ T cells was observed in these draining lymph nodes in COVID-19 patients.

### Effector/memory CD8+ T cells acquire an exhausted phenotype and activated CD4+CTLs may complement CD8+CTLs in late stage COVID-19 lungs

Single cell transcriptomic studies on blood and bronchoalveolar lavage cells in severe COVID-19 has revealed that a large fraction of CD8+ T cells exhibit a dysfunctional gene expression pattern suggestive of exhaustion, with evidence for some retention of functional cells (Wauters et al., 2020; Kusnadi et al. 2021). While PD-1 expression by itself does not necessarily imply a CD8+ T cell is exhausted, a large fraction of the CD8+ T cells in the lungs of patients who succumbed to severe COVID-19 expressed PD-1 (Figures 4A-C). The expression of PD-L1 and IL-27 in the lungs of COVID-19 subjects (Figures S3A and B) represent non-specific findings in an inflammatory context, but their presence is consistent with the possible induction of CD8+ T cell exhaustion (Wherry and Kurachi, 2015). As evidence for recent re-activation, we looked for Granzyme B expression in CD8+ T cells as well as in CD4+ CTLs (Figures 4 D and E). Although only a small fraction of CD8+ T cells express Granzyme B, consistent with the exhaustion phenotype of a large fraction of CD8+ T cells, the absolute numbers of Granzyme B expressing CD4+CTLs and CD8+ T cells were comparable in the resolving group of severe COVID-19 patients (Figure 4F), supporting the view that late in disease, CD4+CTLs may functionally complement CD8+CTLs in this disease.

**Figure 4.**
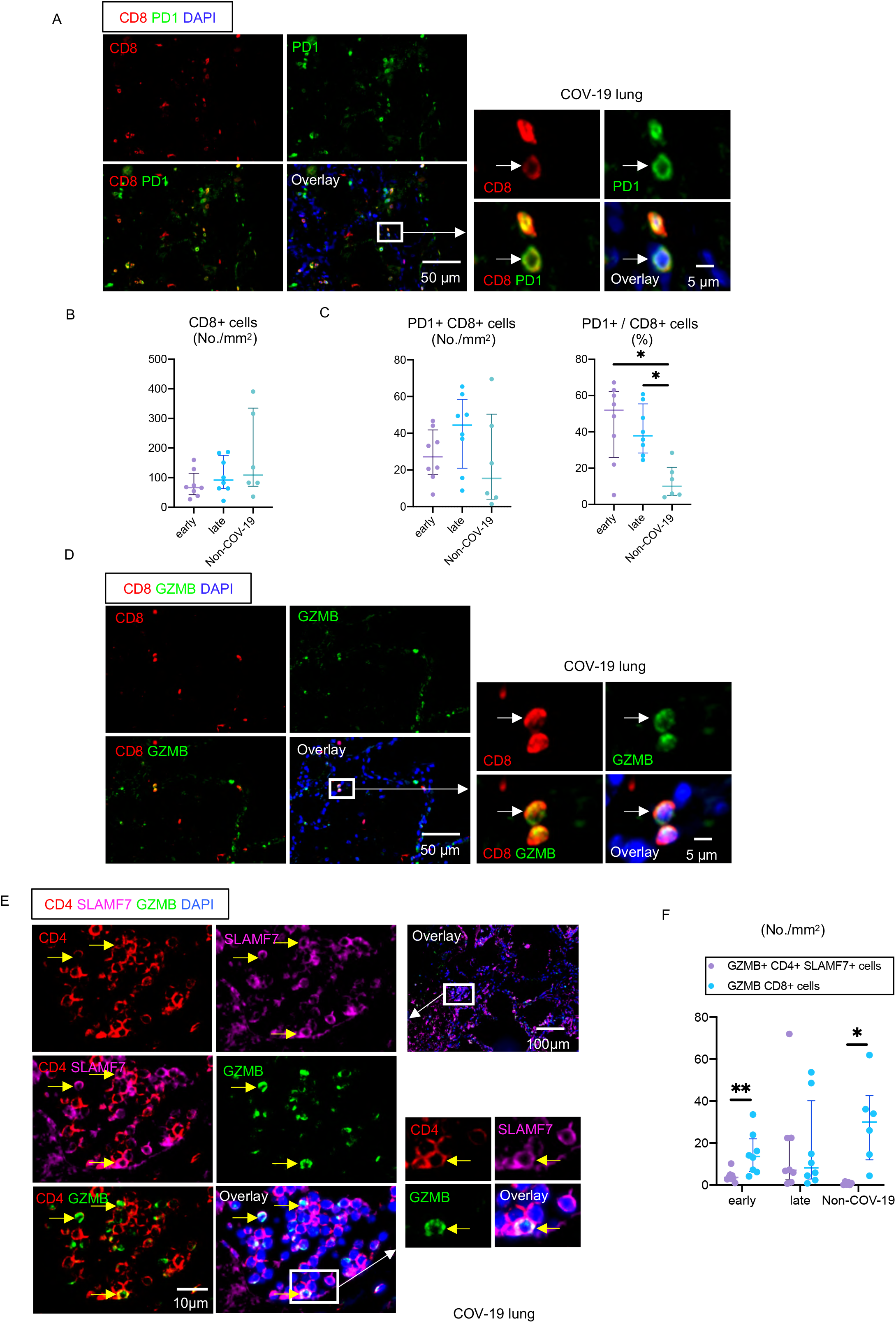
CD8+ T cells infiltrate the lungs but acquire an exhausted phenotype. Representative multi-color immunofluorescence images of CD8 (red), PD1 (green) and DAPI (blue) staining in a lung from a COVID-19 patient. Arrow indicates a PD1+ CD8+ T cell. Absolute numbers of CD8+ T cells in lungs from early (purple) (n = 8) and late (blue) (n = 8) COVID-19 patients and non-COVID-19 patients (green) (n = 6). (C) Absolute numbers and relative proportions of PD1+ CD8+ T cells in lungs from early (purple) (n = 8) and late (blue) (n = 8) COVID-19 patients and non-COVID-19 patients (green) (n = 6). P-values were calculated using the Kruskal-Wallis test to control for multiple comparisons. *p < 0.05. (D) Representative multi-color immunofluorescence images of CD8 (red), GZMB (green) and DAPI (blue) staining in a lung from COV-19 patient. Arrows indicate GZMB expressing CD8+ T cell. (E) Representative multi-color immunofluorescence images of CD4 (red), SLAMF7 (purple), GZMB (green) and DAPI (blue) staining in a lung from a COVID-19 patient. Arrows indicate GZMB+ SLAMF7+ CD4+CTLs. (F) Absolute numbers of GZMB+ CD4+CTLs (purple) and GZMB+ CD8+ T cells (blue) in lungs from early (left) (n = 8) and late (middle) (n = 8) COVID-19 patients and non-COVID-19 patients (right) (n = 6). P-values calculby paired t test. *p < 0.05; **p < 0.01.

### Convergence of the late emergence of CD4+CTLs with increased apoptosis of epithelial cells in severe COVID-19

CD4+ CTLs can contribute to viral clearance (van Leeuwen 2004; Zaunders et al., 2004; Strutt et al., 2014; Weiskopf et al. 2015; Choi et al., 2021) and may be particularly relevant in the context of severe inflammation that results from a fulminant viral infection. CD8+ T cell exhaustion is most likely to occur in such a context to protect the host from catastrophic organ loss. Inflammation also induces HLA class II expression on epithelial and endothelial cells (Wosen et al., 2018). Infected epithelial cells as well as HLA class II expressing macrophages, dendritic cells and B cells that present viral antigen could all be targets of CD4+CTLs. Late in severe COVID-19 about half of all lung epithelial cells express HLA-DR (Figures 5A and B) and epithelial cell apoptosis is particularly prominent in the late subgroup of severe COVID-19 patients (Figures 5C and D).

**Figure 5.**
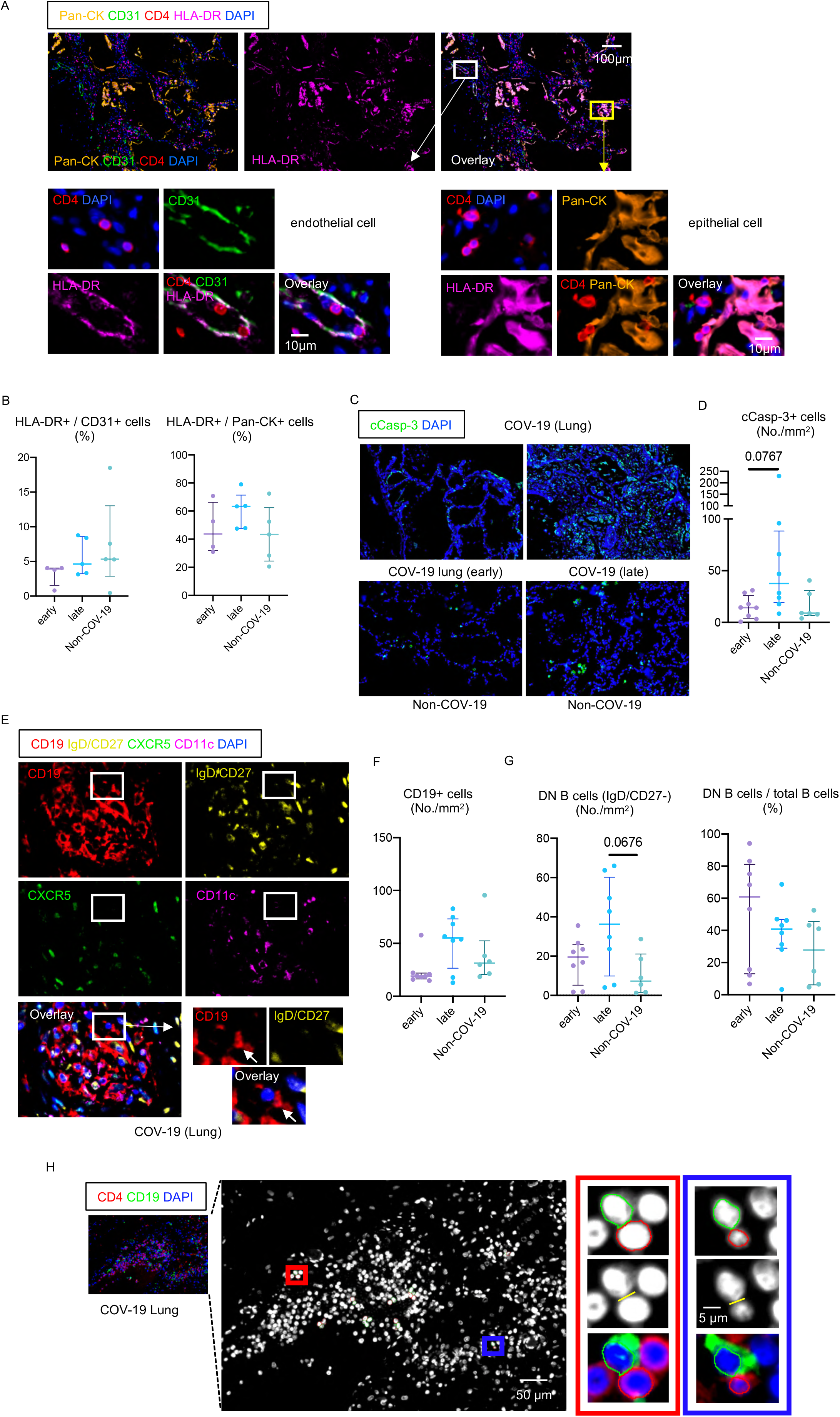
Epithelial cells and endothelial cells express HLA class II and activated B cells accumulate and physically interact with CD4+ T cells in the lungs of COVID-19 patients. (A) Representative multi-color immunofluorescence images of Pan-CK (orange), CD31 (green), CD4 (red), HLA-DR (purple) and DAPI (blue) staining in a lung from a COVID-19 patient. White and yellow boxes highlight HLA-DR+ CD31+ endothelial cells (left) and HLA-DR+ Pan-CK+ epithelial cells (right). CD4+ T cells are observed in close proximity to HLA-DR+ endothelial cells and epithelial cells. (B) Relative proportions of HLA-DR+ endothelial cells (left) and epithelial cells (right) in lungs from early (purple) (n = 4) and late (blue) (n = 5) COVID-19 patients and non-COVID-19 patients (green) (n = 5). (C) Representative multi-color immunofluorescence images of cleaved-caspase3 (c-Casp3) (green) and DAPI (blue) staining in lungs from COVID-19 patients (upper) and non-COVID-19 patients (lower). (D) Absolute numbers of total c-Casp3+ cells in lungs from early (purple) (n = 8) and late (blue) (n = 8) COVID-19 patients and non-COV-19 patients (green) (n = 6). (E) Representative multi-color immunofluorescence images of CD19 (red), IgD/CD27 (yellow), CXCR5 (green), CD11c (purple) and DAPI (blue) staining in a lung from a COVID-19 patient. Arrows indicate IgD/CD27 double negative (DN) B cells. (F) Absolute numbers of total CD19+ B cells in lungs from early (purple) (n = 8) and late (blue) (n = 8) COVID-19 patients and non-COVID-19 patients (green) (n = 6). (G) Absolute numbers and relative proportions of DN B cells in lungs from early (purple) (n = 8) and late (blue) (n = 8) COVID-19 patients and non-COVID-19 patients (green) (n = 6). (H) Representative multi-color immunofluorescence images of CD4 (red), CD19 (green) and DAPI (blue) staining in a lung from a COVID-19 patient. T cells and B cells formed close and extensive intercellular plasma membrane contacts as highlighted in the red and blue boxes. All p-values were calculated using the Kruskal-Wallis test to control for multiple comparisons.

In both rodents and humans, CD4+CTLs turn on a transcriptional program similar to one that is intrinsic to activated CD8+CTLs, but the triggers for CD4+CTL differentiation are not well understood in either species. In human studies, CD4+CTLs can be induced by the activation of naïve CD4+T cells by activated B cells induced to express proteins widely expressed after B cell activation, namely CD70 and OX40L (Choi et al. 2021) though there is no formal genetic evidence for any absolute requirement for B cells or any specific B cell ligands in this differentiation process. In severe COVID-19 we not only noted the presence in the lungs and thoracic lymph nodes of activated B cells, (Figure 5E-G and Figure S3C), but we used cell-cell interaction software to document prominent CD4+T cell-B cell physical interactions in the lung as well (Figure 5H). OX40L is widely expressed in inflamed lymph nodes and an increase in OX40L expressing B cells was observed in COVID-19 lymph nodes (Figure S3E), consistent with a possible contribution of activated B cells to CD4+CTL differentiation.

### CD4+CTLs are expanded in the blood of patients with severe COVID-19

We used 24 color-flow cytometry to analyze effector CD4+ T cell subsets from 62 convalescent and acute COVID-19 patients. CD4+ CD62L^lo^ CD45RA^lo^ effector T cells were gated on and eight major non-overlapping CD4+ T cell subsets (or their close equivalents) were interrogated using surface markers. As seen in Figure 6A, the only CD4+ T cell subsets that were expanded in acute disease compared with convalescence were CD4+ CX3CR1+ (CD4+ CTLs) and CD4+CXCR5-PD-1+ HLA-DR+ (T_PH_) cells. The correlation of CD4+ CTL expansion with disease severity is also depicted in Figure 6B. These data clearly support our findings that CD4+ CTLs represent a prominent and likely functionally relevant CD4+ T cell subset in severe COVID-19

**Figure 6.**
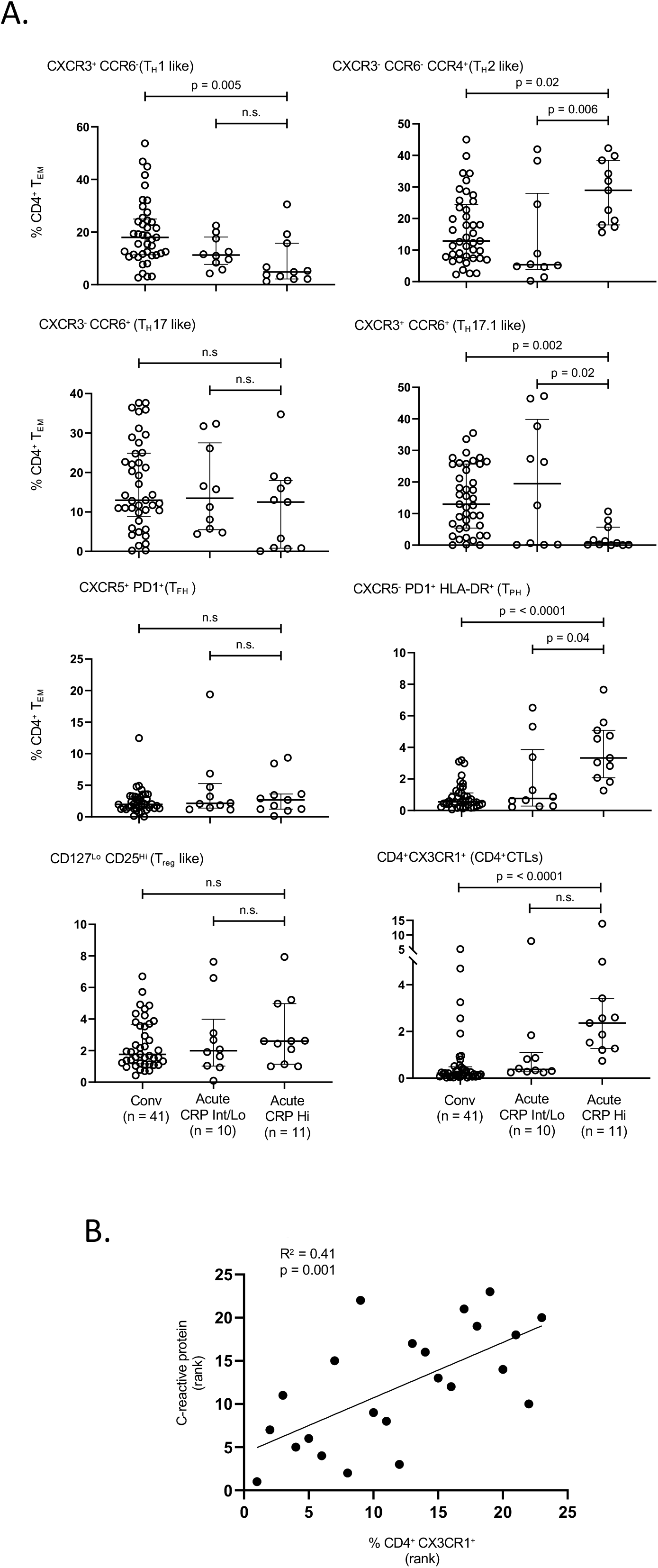
CX3CR1+ CD4+ CTLs expand in the blood with disease severity in severe COVD-19 patients. CD4+ T cells subsets in the peripheral blood of patients with COVID-19 at states of convalescence (n=41), severe illness with an intermediate maximum CRP level during hospitalization (< 200 mg/L; severe (CRP int); n=10), and severe illness with a high maximum CRP level during hospitalization (> 200 mg/L; severe (CRP hi); n=11) as defined by the clinical criteria listed in **Table S3**. A. T_H_1, T_H_2, T_H_17 and T reg cells were defined by surface markers including chemokine receptors and cytokine receptors and not on the basis of transcription factors, hence the nomenclature with a suffix “-like”. Dot plots display circulating T cell subsets as proportions of total CD4_+_ effector-memory T cells. All p-values were calculated using the Kruskal-Wallis test to control for multiple comparisons. CRP = C-reactive protein. Conv = convalescent. B. CD4+CTL accumulation correlates with disease severity as assessed by CRP levels.

These data are consistent with the blood analyses of Meckiff et al. (2020) who examined SARS-CoV-2 peptide-specific CD4+ T cells in COVID-19 using an assay in which the authors gated on activated CD4+ T cells and then performed single-cell RNA seq. While a broadly similar flow cytometry based-activation assay revealed activated CD4+CTLs in the blood that are SARS-CoV-2 specific in COVID-19 patients (Figure S4), we did not pursue this approach further given the broad consonance of our studies on fresh blood samples with the results from Meckiff and colleagues, the potential subset biases that can be introduced by gating strategies in peptide-based T cell activation assays, and the greater relevance we impute to our findings in COVID-19 lungs and draining lymph nodes.

## Discussion

We show here that cytotoxic CD4+ T cells are a prominent CD4+ T cell subset enriched in the lungs in severe COVID-19. The later surge in these cells coincides with a marked increase in pulmonary epithelial cell apoptosis at a stage of disease at which SARS-CoV-2 is known to be depleted in the lungs. Our data suggest that both CD4+CTLs as well as CD8+ T cells likely contribute to the eventual elimination of SARS-CoV-2 infection in the lung, adding SARS-CoV-2 to other viruses such as HIV, dengue and CMV as examples of viral infections in which CD4+ CTLs likely contribute to immune protection (Zaunders et al., 2004; Weiskopf et al. 2015; Strutt et al., 2013; Phetsouphanh et al., 2017). Overall, these data are also consistent with the teleological view that CD8+ T cell exhaustion may have evolved as an organ-protective mechanism during fulminant viral infections and CD4+CTLs may in turn have evolved to complement the cytotoxic functions of CD8+ T cells.

Pulmonary fibrosis was seen in many patients after SARS and MERS (Hui et al., 2005; Zhang et al., 2020b; Das et al., 2017). One of the most detailed follow-up studies in SARS was performed over a 15-year period and involved serial pulmonary CT scans on medical personnel who recovered from the acute disease (Zhang et al., 2020b). 27 of the 71 patients in this study exhibited ground-glass opacities or cord-like consolidations during the follow up period. Unfortunately, one of the long-term sequelae of lung infection with SARS-CoV-2 is the development of lung fibrosis similar to that observed in SARS (Barisione et al., 2020; Grillo et al., 2020; Spagnolo et al., 2020), and the magnitude of this problem is yet to be properly understood. End-stage fibrosis in severe COVID-19 has sometimes required lung transplantation (Bharat et al., 2020).

In other studies, we have implicated CD4+ T cells (and CD8+ T cells) in the induction of apoptotic death and subsequently of fibrosis in IgG4-related disease (Mattoo et al., 2016, Perugino et al., 2020), systemic sclerosis (Maehara et al., 2020), and fibrosing mediastinitis, a disease which is linked to *Histoplasma capsulatum* infection (Allard-Chamard et al. 2021). Others have provided evidence for these cells being of relevance to the fibrosis observed in Grave’s orbitopathy (Wang et al., 2021). In these and other disorders when tissue cells presumably die by apoptosis more rapidly than they can be replenished, the laying down of collagen by activated fibroblasts and myofibroblasts may result in fibrosis. We consider it likely that CD4+CTLs recognizing SARS-CoV-2 peptides presented by HLA class-II molecules probably contributes to the fibrosis seen in some patients with severe COVID-19. This subset of T cells may potentially also be chronically activated by SARS-CoV-2 antigens generated in the intestine for some months after initial infection (Gaebler et al., 2021), or potentially by self-antigen associated with a break in tolerance, and this could possibly contribute to the lung fibrosis identified radiologically by “ground-glass opacities” and seen as part of a broader syndrome that encompass the post-acute sequelae of COVID-19 (PASC), frequently referred to in lay terms as “long COVID”.

No direct and systematic studies on T cell subsets in the lung have previously been reported in severe COVID-19 and as a result no clear picture had so far emerged regarding specific T cell subsets that predominate at a major tissue site of relevance. The only relatively detailed and direct analysis of the lung in severe COVID-19 to date used RNA-ISH and revealed a predominance of inflammatory macrophages in the lung but that study did not provide information on T cell subsets (Desai et al., 2021).

Knowledge of the adaptive immune response in the lung in COVID-19 has so far largely been obtained indirectly and in a non-quantitative fashion by studying bronchoalveolar lavage fluid. The first report that utilized single cell-RNA sequencing of bronchoalveolar exudates in COVID-19 revealed inflammatory monocytes and macrophages that were, in relative terms, more abundant than neutrophils. Some clonal expansion of CD8+ T cells, especially cells with tissue-resident type gene expression was seen but only in mild to moderately ill patients (Liao et al., 2020). In contrast, in severe COVID-19, CD8+ T cells with a naïve-like phenotype predominated. Very little information on CD4+ T cells in the exudates was obtained in this study. In a second study using a broadly similar approach on bronchoalveolar lavage cells, single cell-RNA sequencing was combined with pseudotime inference and T cells in COVID-19 were more exhaustively examined (Wauters et al., 2020). It was inferred that in severe COVID-19, CD8+ T cells decline in relative terms and that both CD8+T cells with a predominantly exhausted-like phenotype and “T_H1_-like” cells exhibit some features of inflammation-induced stress.

In terms of absolute cell numbers, in the absence of COVID-19 the most abundant CD4+ T cells in lymph nodes draining inflamed human lungs are T follicular helper cells. These cells dramatically decline in thoracic lymph nodes in COVID-19 (Figure 3) with a striking loss of Bcl-6 expressing T_FH_ cells (Kaneko et al., 2020; Duan et al, 2020). We speculate that the marked attenuation of Bcl-6 expressing T follicular helper cells reflects a version of an immune deviation phenomenon, wherein activated CD4+ T cells, instead of differentiating into Bcl-6+ T_FH_ cells, are instead directed towards inflammatory fates in the milieu of the draining lymph nodes and primarily differentiate into CD4+CTLs and activated T_H_1 cells. The failure to express Bcl-6 in T cells in a rodent model recapitulates this sort of an immune deviation phenomenon wherein CD4+ T cells chose a more pro-inflammatory fate (Alterauge et al., 2020). High levels of PD-1 in the absence of CXCR5 are found on many activated T_H_1 and T_H_17 cells in lymph nodes, so we chose not to categorize T_PH_ cells (Rao et al., 2017) in tissue sites. However, some of these PD-1 high cells may represent a portion of the extrafollicular and follicular cells that drive robust class switching outside germinal centers and these cells should be investigated in future studies.

In summary, these data indicate that roughly equivalent numbers of activated CD4+CTLs and likely partly exhausted CD8+ T cells are present in the lungs during the resolving phase of severe COVID-19. These cell types are prominent in the adaptive immune landscape and likely contribute both to the clearance of SARS-CoV-2 and are also potential drivers of eventual fibrosis. While initially much of the inflammation in severe COVID-19 may be induced by PAMPs and DAMPs, independent of any T cell contribution, a range of T cells likely contributes to sustaining lung inflammation at the resolving stage of the disease.

### Limitations of the Study

Of necessity, the numbers for the autopsies of non-immunosuppressed patients from whom matched lungs and thoracic lymph nodes were studied were relatively small. While we studied rapidly conducted autopsies with preservation of tissue morphology, autopsy studies are by nature descriptive. Although numerous previous studies have shown the functional ability of CD4+CTLs to kill infected targets, these have all been on the blood, and we could not conduct cytotoxicity studies in the course of this project. We used a single approach to study lymphocyte populations in the lymph nodes and the spleen, and while the complementary use of an orthogonal approach would have been ideal, our data were broadly validated by an analysis of blood lymphocytes by us and by others who have analyzed antigen-specific CD4+ T cells in the blood in severe COVID-19

## Data Availability

There is no genomic or transcriptomic data and there was no clinical trial

## Acknowledgments

We thank members of the Massachusetts Consortium on Pathogen Readiness Specimen Working Group for blood samples. We thank Doug Kwon and Brooke Spencer of the Ragon Institute for access to Tissue Core specimens that were used to validate some of our reagents. This work was supported by NIH U19 AI110495 to SP. Funding for these studies from the Massachusetts Consortium of Pathogen Readiness, the Mark and Lisa Schwartz Foundation and Enid Schwartz is also acknowledged. The graphical abstract was prepared using Biorender.

## Legends to Supplementary Figures

**Figure S1.**
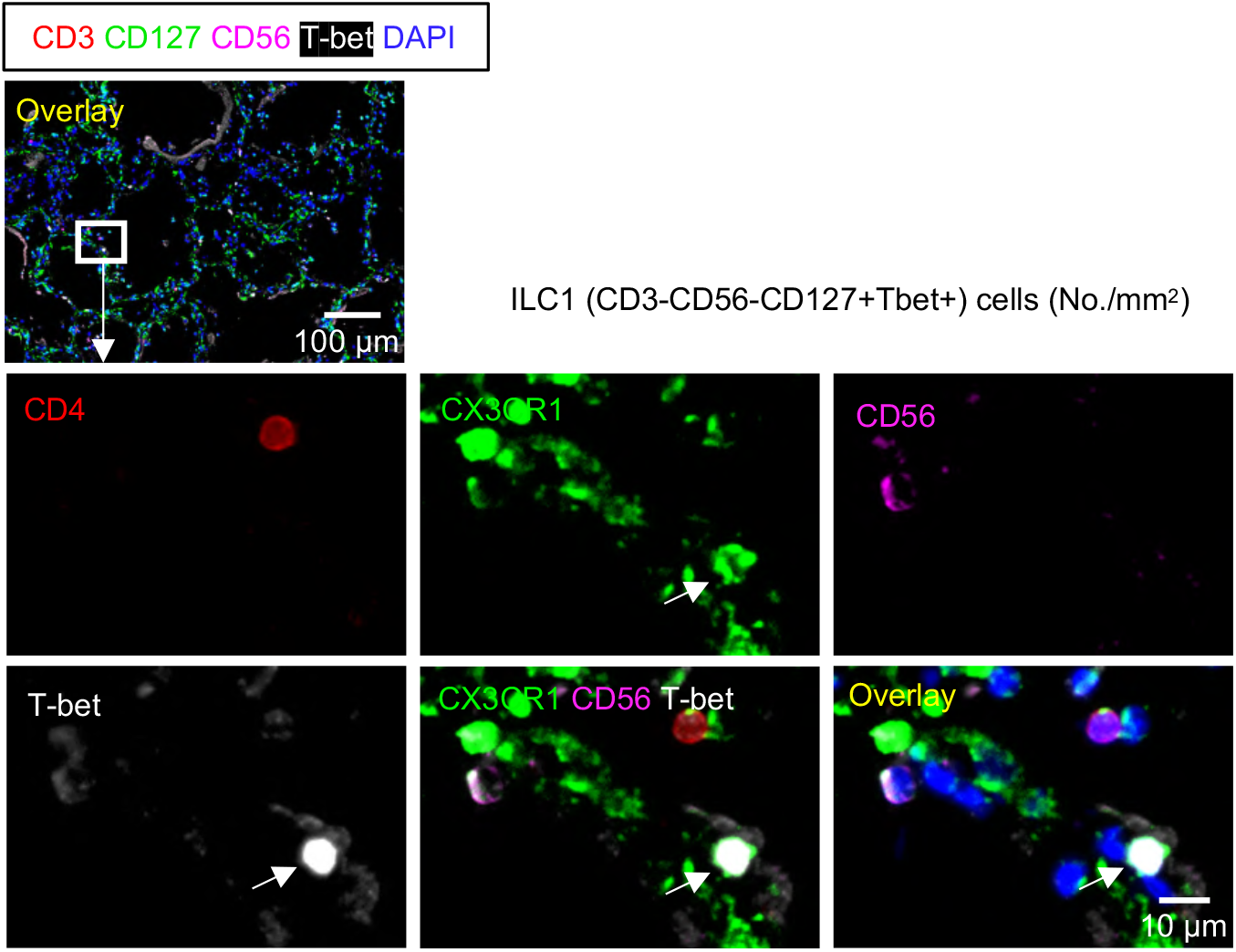
ILC 1s are not prominent in the lungs. Related to Figure 1 Representative multi-color immunofluorescence images of CD3 (red), CD127 (green), CD56 (orange), T-bet (white) and DAPI (blue) staining in a lung from a COVID-19 patient. Arrow indicates a CD3-CD127+ CD56-T-bet+ ILC1.

**Figure S2.**
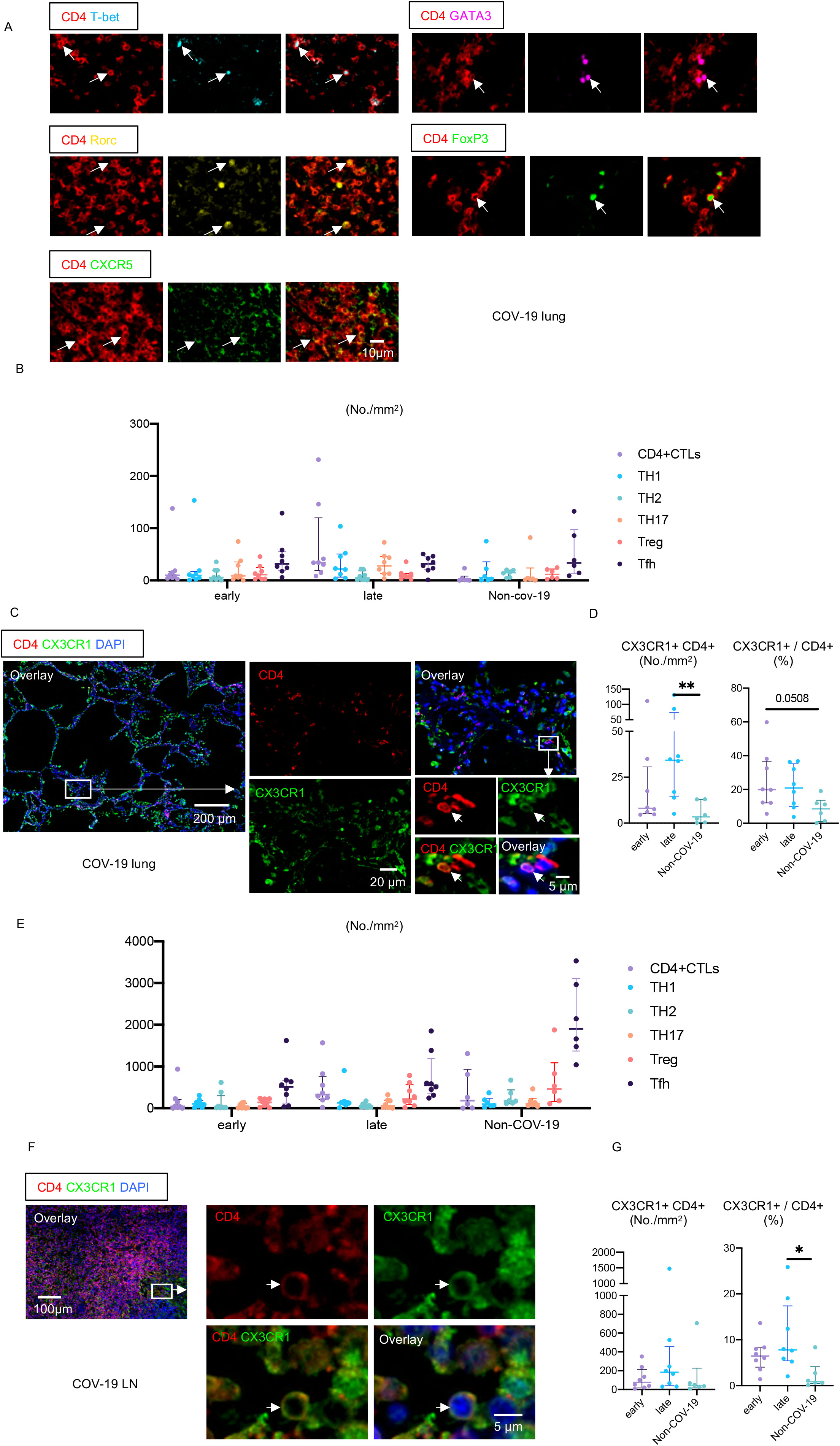
CD4+CTLs are a prominent population in COVID-19. Related to Figures 2 and 3. (A) Representative multi-color staining showing T_H_1, T_H_2, T_H_17, T_FH_ and T _reg_ cells in lungs from COVID-19 patients. [T_H_1: CD4+ (red) T-bet+ (light blue)] [T_H_2: CD4+ (red) GATA3+ (purple)] [T_H_17: CD4+ (red) RORγ+ (yellow)] [T_FH_: CD4+ (red) CXCR5+ (green)] [T_reg_: CD4+ (red) FoxP3+ (green)]. (B) Absolute numbers of all CD4+ T cell subsets in lungs from early (left) (n = 8) and late (middle) (n = 8) COVID-19 patients and non-COVID-19 patients (right) (n = 6). (C) Representative multi-color immunofluorescence images of CD4 (red), CX3CR1 (green) and DAPI (blue) staining in a lung from a COVID-19 patient. (D) Absolute numbers and relative proportions of CX3CR1+ CD4+CTLs in lungs from early (purple) (n = 8) and late (blue) (n = 8) COVID-19 patients and non-COVID-19 patients (green) (n = 6). (E) Absolute numbers of all CD4+ T cell subsets in lymph nodes from early (left) (n = 8) and late (middle) (n = 8) COVID-19 patients and non-COVID-19 patients (right) (n = 6). (F) Representative multi-color immunofluorescence images of CD4 (red), CX3CR1 (green) and DAPI (blue) staining in a lymph node from a COVID-19 patient. (G) Absolute numbers and relative proportions of CX3CR1+ CD4+CTLs in lymph nodes from early (purple) (n = 8) and late (blue) (n = 8) COVID-19 patients and non-COVID-19 patients (green) (n = 6 All p-values were calculated using the Kruskal-Wallis test to control for multiple comparisons. *p < 0.05; **p < 0.01.

**Figure S3.**
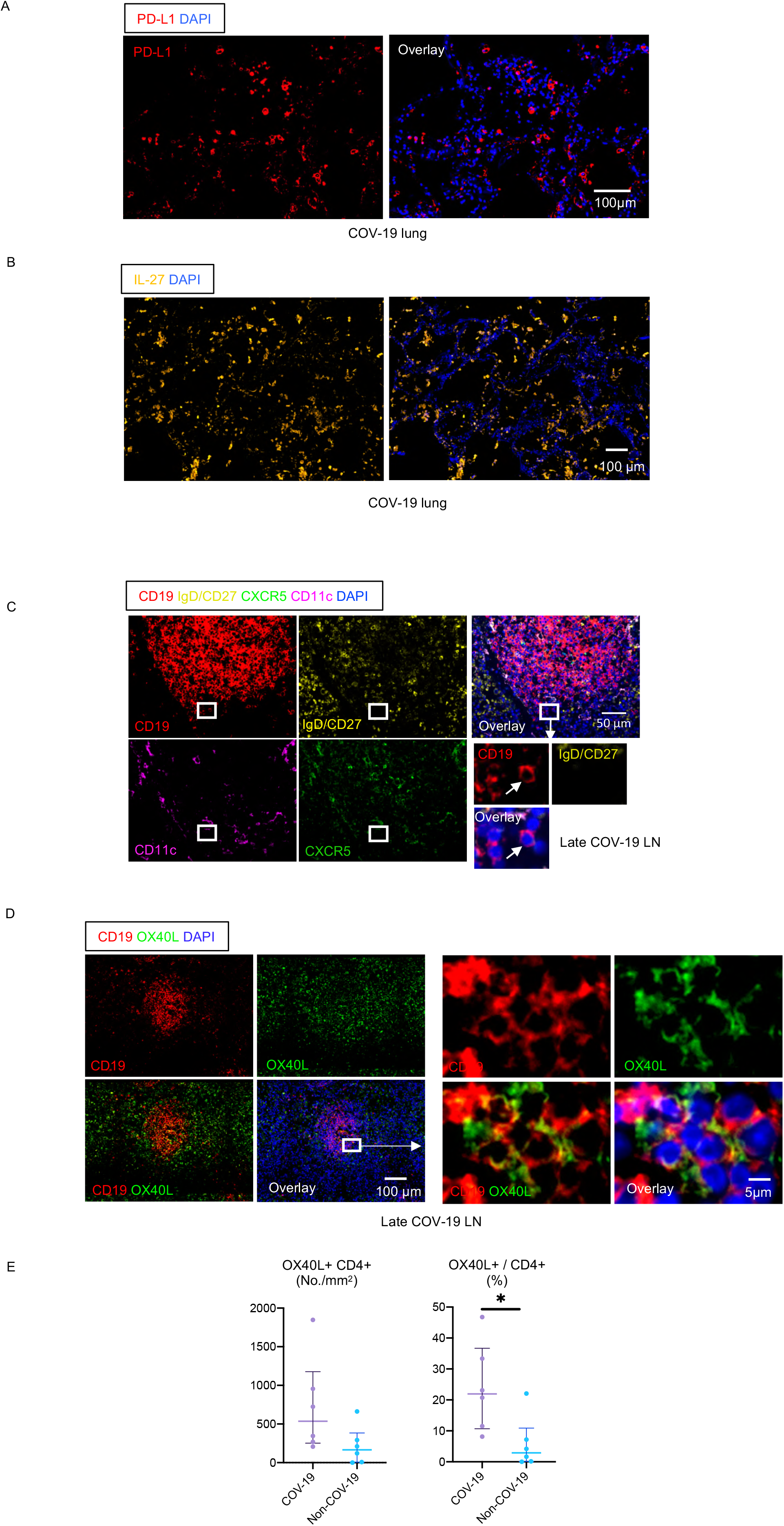
PD-L1 and IL-27 expression and activated B cells in COVID-19 lungs. Related Figure 5. (A) Representative multi-color immunofluorescence images of PD-L1 (red) and DAPI (blue) staining in a lung from a COVID-19 patient. (B) Representative multi-color immunofluorescence images of IL-27 (orange) and DAPI (blue) staining in a lung from a COVID-19 patient. (C) Representative multi-color immunofluorescence images of CD19 (red), IgD/CD27 (yellow), CXCR5 (green), CD11c (purple) and DAPI (blue) staining in a lymph node from a COVID-19 patient. Arrows indicate IgD/CD27 double negative (DN) B cells. (D) Representative multi-color immunofluorescence images of CD19 (red), OX40L (green) and DAPI (blue) staining in a lung from a COVID-19 patient. (E) Absolute numbers and relative proportions of CX3CR1+ CD4+CTLs in lymph nodes from COVID-19 patients (purple) (n = 6) and non-COVID-19 patients (blue) (n = 6). P-values were calculated by the Mann-Whitney *U* test. *p < 0.05.

**Figure S4.**
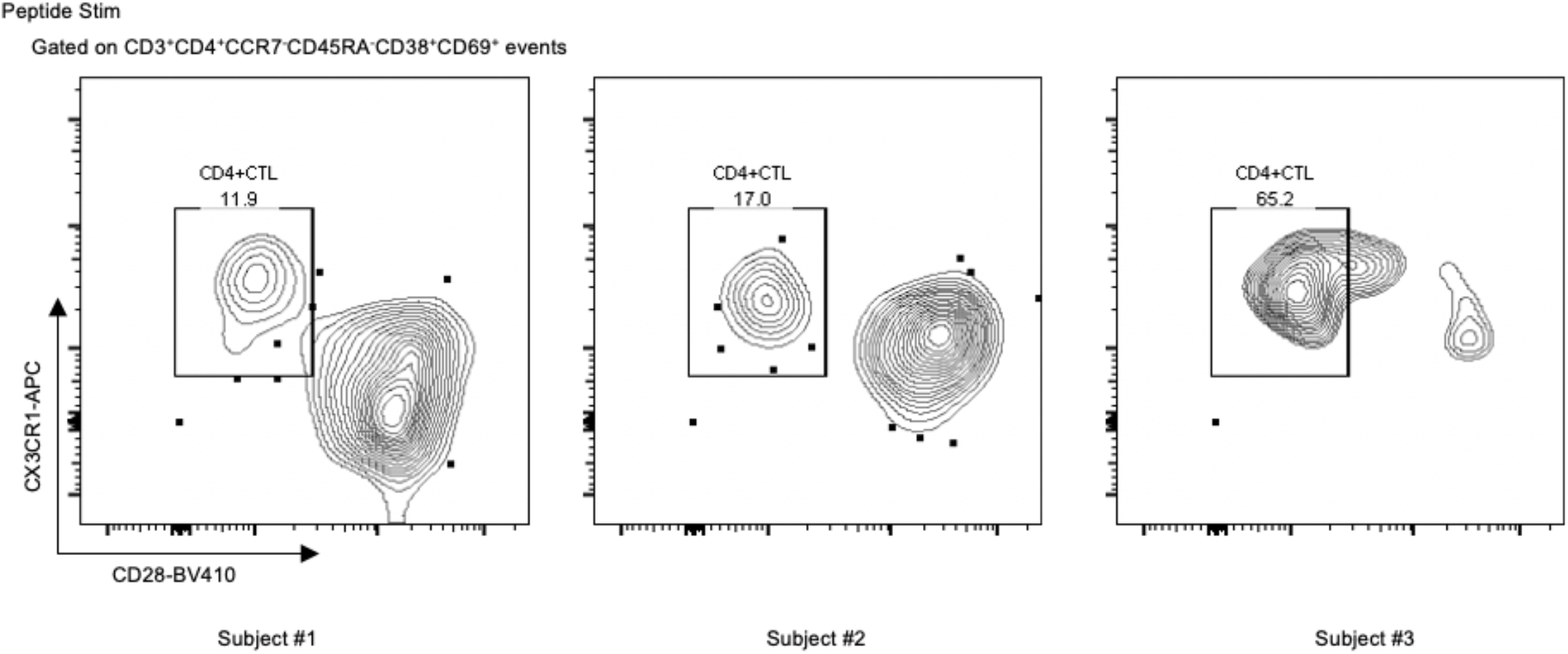
SARS-CoV-2 peptides induce the activation of CD4+ CTLs from COVID-19 patients. Related to Figure 6. SARS-COV-2 peptides were incubated overnight with PBMCs from COVID-19 patients and then within the CD3+CD4+CCR7-CD45RA-gate, the enrichment of CX3CR1+CD28lo CD4+ CD4+CTLs among CD38+CD69+ stimulated cells as compared to CD38-CD69-unstimulated cells was compared.

## Human Subjects

### Tissue analysis Cohort

Thoracic lymph nodes and lung samples from COVID-19 patients were obtained through the Brigham and Women’s Hospital Department of Pathology. Patients who had been on trials for known immunosuppressive agents were excluded. Controls were age- and gender-matched individuals with lung disease who were exhaustively investigated when hospitalized but had no evidence for SARS-CoV-2 infection (Table S2; Kaneko et al., 2020)

#### Sample Size Estimation

Power calculations and sample size estimation were not performed before the initiation of this study which was based primarily on the availability of COVID-19 positive autopsies for analysis in 2020. All cases were retrieved from the Anatomic Pathology files of Brigham and Women’s Hospital and included 22 patients (information on age and gender in Table S1 and S2) including 16 with laboratory confirmed COVID-19 who underwent autopsy in 2020.

#### Sample allocation to experimental groups

All patients had tested positive for SARS-CoV-2 by RT-PCR of nasopharyngeal swabs in a laboratory during hospital admission. All cases were divided into two groups; early (less than ten days from respiratory symptoms onsets to death, hospitalization of up to 8 days), and late (hospitalized for 15-36 days prior to death). Thoracic lymph nodes were obtained from six age-matched non-COVID-19 patients who underwent autopsies at the Brigham and Women’s Hospital in the same time window.

### Peripheral Blood Cohort

Peripheral blood samples were drawn from both Outpatients and Inpatients with COVID-19 at Massachusetts General Hospital and fresh blood was analyzed for flow cytometry using a multi-color panel.

Sample Size estimation: Power calculations were not undertaken prior to the initiation of these studies. However, the primary end-point was the sum of activated T cells (as a % of CD4+ T cells in peripheral blood). Using a two-sided Student’s t-test to compare log values of convalescent and severe (CRP hi) COVID-19 patient samples, with groups of 10, we had more than 90% power to detect an effect of size of 1.20 between groups based on simulation studies using 10,000 Monte Carlo samples with a type 1 error rate of 5%.

Allocation to Experimental Groups: Data is presented on T cell populations from 62 patients, including moderately ill, severely ill and convalescent patients. Convalescence was defined as a clinically asymptomatic state on the date of blood draw, either from a baseline asymptomatic state or recuperated from moderate clinical symptoms of COVID-19. Moderate disease was defined as active clinical symptoms of COVID-19 on the date of blood draw that did not necessitate a hospital admission. Severe disease was defined as active clinical symptoms of COVID-19 on the date of blood draw that did necessitate a hospital admission. Severe disease was further subdivided by maximum CRP level during hospital admission as CRP < 200 mg/L classified as ‘CRP intermediate (int)’ and CRP > 200 mg/L as ‘CRP high (hi)’.

### Study Approval

This study was performed with the approval of the Institutional Review Boards at the Massachusetts General Hospital and the Brigham and Women’s Hospital.

## METHOD DETAILS

### Multi-color immunofluorescence staining

Tissue samples were fixed in formalin, embedded in paraffin, and sectioned. These specimens were incubated with the following antibodies: anti-CD3 (A045229-2; DAKO), anti-CD4 (ab133616; Abcam), anti-CD19 (SKU310; Biocare Medical), anti-Pan-CK (ab27988; Abcam), anti-CD31 (3528; Cell Signaling Technology), anti-citrullinated histone 3(ab5103; Abcam), anti-MPO (ab9535; Abcam), anti-CD68 (ab955; Abcam), anti-ASC1 (13833S; Cell Signaling Technology), anti-CD56 (99746; Cell Signaling Technology), anti-CD127 (B2830; Lifespan Bioscience), anti-IgD (AA093; DAKO), anti-CD27 (ab131254; Abcam), anti-CD11c (ab52632; Abcam), anti-SLAMF7 (HPA055945; Sigma-Aldrich), anti-CX3CR1 (ab8021; Abcam), anti-T-bet (ab150440; Abcam), anti-GATA3 (MA1028; Invitrogen), anti-Ror gamma (ab212496; Abcam), anti-CXCR5 (clone: MAB190; R&D Systems), anti-FoxP3 (clone: 98377; Cell Signaling Technology), anti-CD8 (ab85792; Abcam), anti-PD1 (B13300; Lifespan Bioscience), anti-PD-L1 (13684; Cell Signaling Technology), anti-GZMB (ab4095; Abcam), anti-HLA-DR (ab20181; Abcam), anti-IL27 (B2719; Lifespan Bioscience), anti-OX40L (B10083; Lifespan Bioscience) and anti-cleaved caspase-3 (9664; Cell Signaling Technology) followed by incubation with a secondary antibody using an Opal™ Multiplex Kit (Perkin Elmer). The samples were mounted with ProLong™ Diamond Antifade mountant containing DAPI (Invitrogen).

### Microscopy and Quantitative Image Analysis

Images of the tissue specimens were acquired using the TissueFAXS platform (TissueGnostics). For quantitative analysis, the entire area of the tissue was acquired as a digital grayscale image in five channels with filter settings for FITC, Cy3, Cy5 and AF75 in addition to DAPI. Cells of a given phenotype were identified and quantitated using the TissueQuest software (TissueGnostics), with cut-off values determined relative to the positive controls. This microscopy-based multicolor tissue cytometry software permits multicolor analysis of single cells within tissue sections similar to flow cytometry. StrataQuest (TissueGnostics) software was also used to quantify cell-to-cell contact. In the StrataQuest cell-to-cell contact application, masks of the nuclei based on DAPI staining establish the inner boundary of the cytoplasm and the software “looks” outwards towards the plasma membrane boundary. Overlap of at least 3 pixels of adjacent cell markers is required to establish a “contact” criterion. Although the software has been developed and validated more recently, the principle of the method and the algorithms used have been described in detail elsewhere (Ecker and Steiner, 2004)

### Quantification and statistical analysis

For tissue studies, flow cytometry and clinical correlations, data are shown as median [interquartile range: 25^th^-75^th^ percentiles]. GraphPad Prism version 8 was used for statistical analysis, curve fitting and linear regression. A two-tailed Mann-Whitney U test was used to calculate p values for continuous, non-parametric variables. The paired t test was also performed to calculate p-values between two variables in the same subject. For comparing more than one population, Kruskal-Wallis testing was used with Dunn’s multiple comparison testing. A p value of < 0.05 was considered significant.

### Flow cytometry

1 Million fresh PBMCs were stained within 2 hours of isolation. Prior to antibody staining, Fc receptors were blocked using human FcR blocking reagent (Miltenyi) at a concentration of 1:50 at 4 °C for 10 minutes. Cells were surface stained at 4 °C, protected from light, using optimized concentrations of fluorochrome-conjugated primary antibodies for 30 minutes as well as live/dead fixable blue stain (Thermo Fisher) at a concentration of 1:20 using the following antibody panels (clone, manufacturer): CD14 (HCD14, Biolegend), CD19 (HIB19, Biolegend), CD34 (581, Biolegend), CD94 (HP-3D9, BD Biosciences), CD11c (Bu15, Biolegend), CD3 (UCHT1, BD Biosciences), CD4 (SK3, BD Biosciences), CD8 (RPA-T8, BD Biosciences), CD38 (HIT2, BD Biosciences), HLA-DR (L243, Biolegend), CD57 (NK-1, BD Biosciences), CD25 (2A3, BD Biosciences), CXCR3 (G025H7, Biolegend), CCR6 (G034E3, Biolegend), CCR4 (L291H4, Biolegend), CX3CR1 (2A9-1, Biolegend), PD-1 (EH12.1, BD Biosciences), CXCR5 (RF8B2, BD Biosciences), CD62L (DREG-56, Biolegend), CD45RA (HI100, BD Biosciences), CD127 (A019D5, Biolegend), CD16 (3G8, BD Biosciences), CD56 (HCD56, Biolegend).

Cells were then washed in PBS and fixed using 4% paraformaldehyde for 30 minutes at at 4. Flow cytometry was performed on a BD Symphony (BD Biosciences, San Jose, CA) and rainbow tracking beads (8 peaks calibration beads, Fisher) were used to ensure consistent signals between flow cytometry batches. FCS files were analyzed, and B cell subsets were quantified using FlowJo software (version 10).

### Peptide Stimulation of PBMCs

PBMCs from 3 hospitalized subjects with acute COVID-19 were used for the activation-induced marker assay. Briefly, cryopreserved PBMCs were thawed and rested at 37°C prior to peptide stimulation. Cells were quantified and plated at a concentration of approximately 2.5 million cells per mL in AIM-V media. Cells were then incubated for 16 hours at 37C, 5% CO_2_ with or without the addition of an overlapping peptide pool of Spike antigen (JPT Peptide Technologies; PM-WCPV-S) according to the manufacturer’s recommended protocol. Cells were harvested, washed, and underwent standard Fc block and cell staining for flow cytometric analysis. Additional antibodies used in this assay not included in the other blood studies included mouse anti-human CD69-BB700 (BD Bioscience, clone FN50, stained at 1:50 at 4C), mouse anti-human CD38-BUV661 (BD Bioscience, clone HIT2, stained at 1:400 at 37C), mouse anti-human CX3CR1-APC (BioLegend, clone 2A9-1, stained at 1:50 at 4C), and mouse anti-human CD28-BV480 (BD Bioscience, clone CD28.2, stained at 1:100 at 37C)

### Study Approval

This study was conducted with the approval of the Institutional Review Boards at the Massachusetts General Hospital and the Brigham and Women’s Hospital.

## List of Supplementary Tables

Table S1. Autopsied COVID-19 Patients studied by immunofluorescence and multispectral imaging (Related to Figures 1-5)

Table S2. Non-COVID-19 Autopsied Patients (Related to Figures 1-5)

Table S3. Summary of Information on COVID-19 patients in whom circulating T cells were analyzed by flow cytometry (Related to Figure 6)

---

**Table S1.** Autopsied COVID-19 Patients studied by immunofluorescence and multispectral imaging.

| Group | early | Late (resolving) |
| --- | --- | --- |
| Numbers | n=8 | n=8 |
| Duration of illness | <10 days | 15-36 days |
| Demographics |  |  |
| Age (years) (mean ± s.d.) | 69.5 (±14.4) | 66.6 (±9.1) |
| Gender (% male) | 87.5% | 50% |

**Table S2.** Non-COVID-19 Autopsied Controls Age range 64-78 (Male 67%, Female 33%)

| **Presumed Causes of Death** |
| --- |
| Multi-organ system failure, pancreatitis |
| Non-metastatic bladder malignancy |
| Multi-organ system failure, thromboembolic infarcts |
| Respiratory failure, treatment resistant MAC |
| Cerebellar hemorrhage |
| Fibrinous pericarditis, subpleural fibrosis |

**Table S3.** Blood Studies on Acute and Convalescent COVID-19 patients.

| **Convalescent** | **Acute CRP int/lo** | **Scute CRP high** |
| --- | --- | --- |
| **n=39** | **n= 10** | **n=11** |
| **CRP - na** | **CRP 107.1(IQR 67.5-149.4)** | **300 (IQR 279.3-338.4)** |

Convalescence was defined as a clinically asymptomatic state on the date of blood draw, either from a baseline asymptomatic state or recuperated from moderate clinical symptoms of COVID-19.
Severe disease was further subdivided by maximum CRP level during hospital admission as CRP < 200 mg/L 'Acute CRP int/lo' and CRP > 200 mg/L 'Acute CRP high.'

